# Heritability of Alzheimer-related plasma biomarkers in the Amish population

**DOI:** 10.1101/2025.05.13.25327557

**Authors:** Ping Wang, Yeunjoo E. Song, Audrey Lynn, Kristy Miskimen, Alex Gulyayev, Michael B. Prough, Daniel A. Dorfsman, Renee A. Laux, Sarada L. Fuzzell, Sherri D. Hochstetler, Andrew F. Zaman, Larry D. Adams, Laura J. Caywood, Jason E. Clouse, Sharlene D. Herington, Patrice Whitehead, Yining Liu, Noel Moore, Paula Ogrocki, Alan J. Lerner, Anthony J. Griswold, Jeffery M. Vance, Michael L. Cuccaro, William K. Scott, Margaret A. Pericak-Vance, Jonathan L. Haines

## Abstract

**INTRODUCTION:** Plasma biomarkers for Alzheimer disease (AD) hold promise for disease diagnosis and prediction, yet their genetic underpinnings remain underexplored.

**METHODS:** We measured plasma amyloid beta 40 (Aβ40), Aβ42, Aβ42/Aβ40, total tau (t-tau), phosphorylated tau 181 (p-tau181), Aβ42/t-tau, Aβ42/p-tau181, neurofilament light chain, and glial fibrillary acidic protein in the Midwestern Amish. Pedigree-based heritability 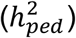 was estimated from multigenerational pedigrees, and SNP-based heritability 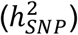 was derived from single nucleotide polymorphisms (SNPs).

**RESULTS:** Among all Amish individuals, additive genetic effects 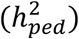 explained 11.1% to 36.6% of biomarker variances. 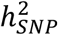 estimates were consistently lower, ranging from 6.7% to 28.7%. The heritability of these biomarkers in subgroups of cognitively normal individuals and *APOE* ε4 non-carriers yielded similar results.

**DISCUSSION:** Plasma biomarkers such as amyloid β, t-tau, and p-tau181 are moderately heritable in the Amish, underscoring the impact of genetic determinants of plasma biomarkers associated with AD.

## 1. Background

Alzheimer disease (AD) is the most common cause of dementia, marked by progressive deficits in memory, cognition, language, behaviors, and daily function. The complexity of AD arises from the interplay of genetics, environment, and lifestyle factors, with genetics playing a substantial role in shaping both the risk and the progression of AD. The majority of AD cases are sporadic, tend to have late-onset symptoms, and do not have a single genetic cause. Advances in genome-wide association studies (GWAS) have identified over 70 loci associated with late-onset AD (LOAD);^1–5^ however, the genetic architecture of AD is not fully understood. Previous twin studies on prevalent or incident cases estimated the heritability of AD to range between 37% and 79%;^6–8^ however, common variations identified through GWAS explain only 12% - 57% of this heritability, with apolipoprotein E (*APOE*) estimated to contribute up to 27% as a major risk factor.^9,10^ This gap, often referred to as the “missing heritability,” underscores the need for further exploration of genetic factors for AD.^11^

Recent efforts, such as studies on gene-environment and gene-gene interactions,^12–15^ along with large-scale multi-ancestry GWAS,^2,3,5,16^ have improved the power to explain more heritability. However, these strategies primarily rely on samples with binary classifications of clinical AD or AD-by-proxy, which introduces several limitations. Because of the diverse phenotypes, clinical diagnosis of biological AD may have misdiagnosis rates of up to 30%.^17^ The reliance on binary classification in genetics research neglects individuals in preclinical or mild cognitive impairment stages who may share risk factors with AD individuals and fails to capture the continuum of AD pathology. This is especially concerning with current trends of genetic discoveries through large-scale GWAS incorporating the AD diagnosis by proxy.^18^ Quantitative endophenotypes offer a promising solution to these challenges by providing an additional path to search for missing heritability of AD.

Biomarkers are endophenotypes that correlate with disease liability and pathological changes. In AD, the aggregates of amyloid beta (Aβ) plaques and tau tangles in the brain are the primary pathological hallmarks.^19,20^ Recently, neurofilament light chain (NfL) has emerged as a general biomarker for neuroaxonal damage and degeneration,^21^ and glial fibrillary acidic protein (GFAP) reflects neuroinflammation and astrocytic activation,^22^ both are processes central to AD pathogenesis. Although these biomarkers can be reliably quantified *in vivo* using neuroimaging and/or cerebrospinal fluid measures, such collection methods are invasive, expensive, and infeasible for large-scale population studies. In contrast, plasma biomarkers offer a promising solution, with recent technological advances enabling the detection of extremely low levels in the blood with high sensitivity.^23,24^ Plasma biomarkers can be measured in all individuals at all AD stages and can provide greater power than binary diagnosis to assess the genetic contribution related to AD.

Despite their potential, the heritability of AD-related plasma biomarkers remains underexplored. Early work using pedigrees in LOAD families estimated the heritability of plasma Aβ40 and Aβ42 at 56.1% and 40.7%, respectively.^25^ More recent twin studies further calculated the heritability of several plasma biomarkers, with values ranging from 16% for the Aβ42/Aβ40 ratio to as high as 60% for GFAP.^26,27^ While these findings highlight a genetic influence on plasma biomarkers, variability exists, and key biomarkers like phosphorylated tau (p-tau181) have not been evaluated.

To address this gap, we estimated the heritability of plasma Aβ40, Aβ42, Aβ42/Aβ40, total tau, p-tau181, Aβ42/t-tau, Aβ42/p-tau181, NfL, and GFAP levels in the Midwestern Amish, using their extensive pedigree information and individual-level genotype data. The Amish are ideal for genetic research due to their unique genetic and cultural background as a founder population.^28^ The extended multigenerational families and relatively uniform lifestyles in the Amish provide a powerful tool to estimate the heritability with less influence from the variability in the shared environment.

## 2. Methods

### 2.1 Study population

We analyzed data from the Collaborative Amish Aging and Memory Project (CAAMP), a prospective study investigating successful aging and dementia within the Amish communities of Ohio (OH) and Indiana (IN). The Amish represent an isolated founder population, primarily descended from German and Swiss Anabaptist immigrants who settled in the United States during the 18th and 19th centuries.^29^ Specifically, the Amish living in and around Holmes County (OH), Elkhart County (IN), and LaGrange County (IN) are mostly descendants from the German Palatinate; meanwhile, those in Adams County (IN) trace their ancestry to Swiss Anabaptist immigrants.^29,30^ The recruitment and ascertainment process for the CAAMP study has been previously described.^31,32^ Briefly, a door-to-door interview was employed to gather demographic information, medical histories, blood samples, and cognitive assessments. An all-connecting pedigree was generated using records from the Anabaptist Genealogy Database,^30^ enabling the determination of kinship among the Amish participants.

### 2.2 Clinical phenotypes

Upon enrollment, all participants underwent cognitive screening. Based on established cutoffs,^32^ a clinical adjudication team of neurologists and neuropsychologists determined a consensus diagnosis for each Amish participant. Individuals were categorized as cognitively unimpaired (CU), having mild cognitive impairments (MCI), cognitively impaired but not AD (CINAD), or cognitively impaired related to AD (AD). A few individuals requiring additional follow-up could not be initially characterized and were excluded from the analysis.

### 2.3 Plasma biomarkers and quality control

Plasma biomarkers were measured from frozen samples stored in 500 μL aliquots at −80°C, using the Quanterix™ HD-X platform. This platform leverages the Simoa™ technology for ultra-sensitive protein detection with a lower limit of 0.1 pg/mL. Specifically, plasma Aβ40 and Aβ42 levels were initially measured using the Simoa™ Neurology 3-Plex A (N3PA) assay, then transitioned to the Neurology 4-Plex E (N4PE) assay following its release in 2017. Plasma total tau levels were measured using the N3PA assay for all samples. Plasma NfL and GFAP levels were assessed using the N4PE assay. Plasma p-tau181 was measured with the Simoa™ P-tau181 Advantage V2 assay. Results for all biomarkers were above the detection limit of the assays.

Initial biomarker data were processed using the Quanterix Analyzer v1.6 software ^33^ to calculate standard curves and quantify plasma protein levels. Quality control (QC) procedures were conducted at the individual biomarker level to ensure the reliability of the data (Supplemental Figure 1). Biomarker samples were matched to clinical data from the Amish population, and those lacking a consensus diagnosis or originating from non-Amish individuals (internal controls) were excluded. For individuals with repeated results from samples collected on the same date, the set with the higher intra-assay coefficient of variation (CV) was excluded. Only samples with a CV below 20% were retained for analysis. Based on summary statistics and visualization, we evaluated samples for each biomarker on a case-by-case basis. The outlier cut-off for each biomarker was defined by the Median Absolute Deviation (MAD). This method is more robust in detecting outliers for skewed distributions due to its reliance on non-parametric measures of central tendency and variation. From biomarkers that passed QC, we restricted our data to the most recently measured biomarkers for this study. Only 1-3 samples fell beyond the outlier cut-off per biomarker.

**Figure 1.**
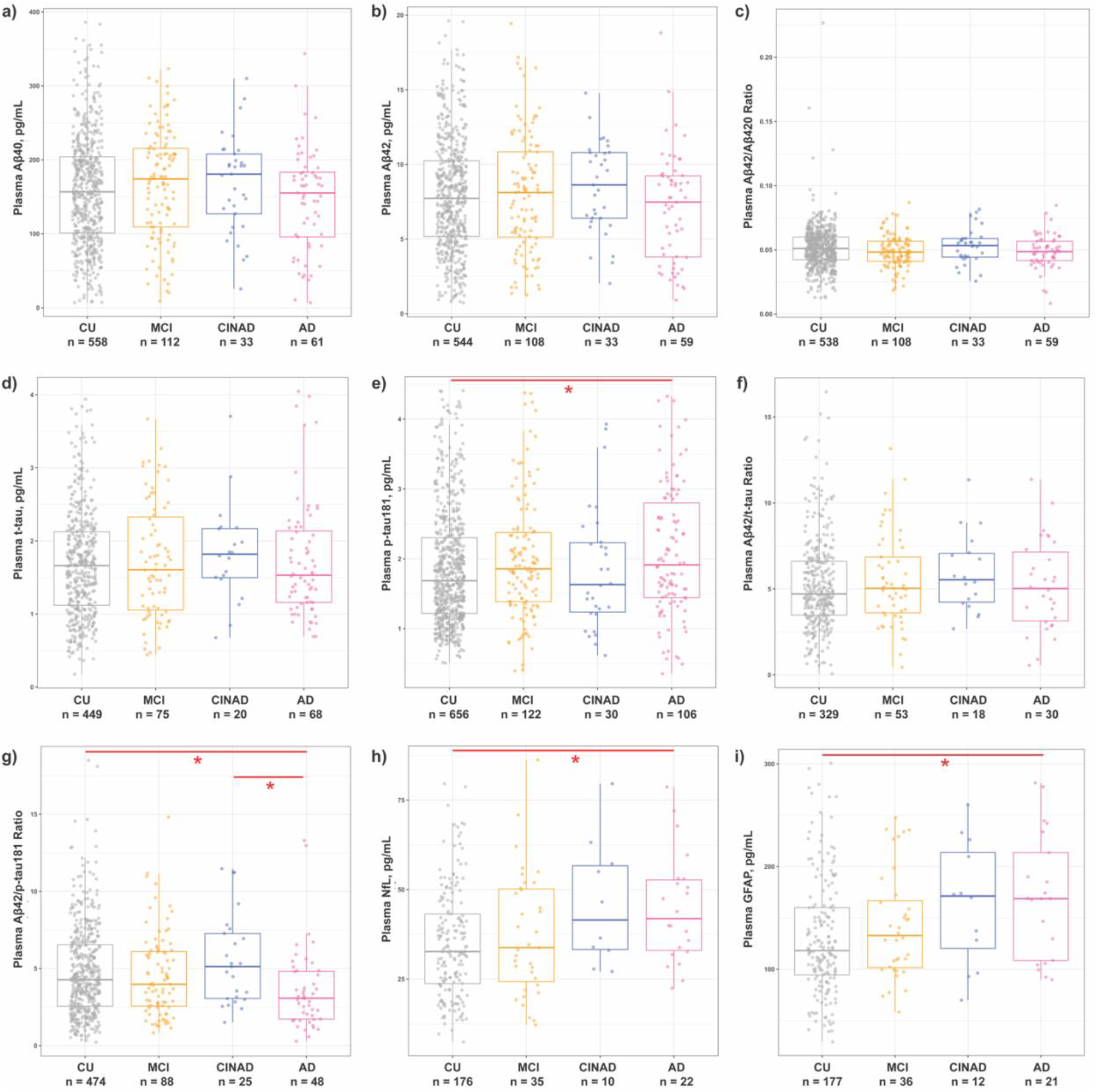
Plasma p-tau181, Aβ42/p-tau181 ratio, NfL, and GFAP levels were significantly different in Amish individuals with AD. Boxplots comparing a) plasma Aβ40 levels; b) plasma Aβ42 levels; c) plasma Aβ42/Aβ40 ratio; d) plasma total tau levels; e) plasma p-tau181 levels; f) plasma Aβ42/t-tau ratio; g) plasma Aβ42/p-tau181 ratio; h) plasma NfL levels; i) plasma GFAP levels across diagnostic groups. The box plots depict the median (horizontal bar), interquartile range (IQR, hinges) and minimal and maximal values (whiskers). * *p* < 0.05 after correcting for multiple comparisons using the Bonferroni method. Abbreviations: CU, cognitively unimpaired; MCI, mild cognitive impairment; AD, Alzheimer disease; CINAD, cognitively impaired but not AD; Aβ, amyloid beta; t-tau, total tau; p-tau181, phosphorylated tau at epitope 181; NfL, neurofilament light chain; GFAP, glial fibrillary acidic protein.

### 2.4 Genotyping, imputation, and quality control

30 mL of blood was collected from Amish individuals for DNA extraction. Genotyping was conducted using either the Illumina Expanded Multi-Ethnic Genotyping Array (MEGAEX) with custom content ^34^ or the Illumina Global Screening Array (GSA).^35^ The MEGAEX chip includes over 2 million markers, and the GSA chip includes a base quantity of 660,000 markers. To capture novel variants in the Amish, about 6,000 more variants are added to the GSA chip genotyping, including over 1,100 novel variants that were identified from previous whole exome or whole genome sequencing studies in the Amish^31,32,36,37^ and other AD-related variants from the Alzheimer’s Disease Sequencing Project.^38^

Imputation was carried out using the Michigan Imputation Server and the Trans-Omics for Precision Medicine (TOPMed) panel.^39^ The MEGAEX and GSA array data were imputed separately, using the GRCh38 build for autosomes and hg19 for the X chromosome. Following imputation, downstream QC was performed independently on each dataset prior to merging. QC procedures were implemented on the merged genotype data using PLINK 2.0 and included evaluations of family relationships, sex discrepancies, and batch effects. Single nucleotide polymorphisms (SNPs) were filtered based on low call rates (missing > 5%), minor allele frequency (MAF < 0.05), and Hardy-Weinberg equilibrium (HWE < 1e-6). Ultimately, 4,403,280 SNPs passed the QC across both genotyping platforms. The QC process was repeated for biomarker-specific datasets prior to heritability estimation.

### 2.5 Statistical analysis

For group comparisons of continuous variables with non-normal distributions, the Kruskal-Wallis test was employed. Categorical variables were analyzed using the Chi-square test. We applied the Bonferroni method to correct for multiple comparisons (n = 6 tests) in the post-hoc analysis.

A prior analysis of Amish genealogy established a large 14-generation pedigree encompassing all Amish individuals.^40^ For this study, individuals with biomarkers that passed QC were organized into smaller pedigrees based on shared grandparental lineage. Each biomarker was standardized using a rank-based inverse normal transformation after QC. Pedigree-based heritability 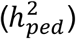 was estimated using a variance component model implemented in the Sequential Oligogenic Linkage Analysis Routines (SOLAR) packages,^41^ adjusting for age, sex, the presence/absence of *APOE* ε4 alleles, diagnosis, study center, and biomarker batches. SOLAR utilizes a maximum likelihood estimate to analyze pedigrees of arbitrary size and is designed to handle discrete and quantitative traits with covariate adjustments. For family-based quantitative data, SOLAR decomposes the phenotypic variance into genetic and environmental components based on the observed covariance in a trait among family members. The statistical significance of 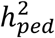 was determined by a likelihood ratio test, comparing the log-likelihood of the estimated model to a null model where the additive genetic component was fixed to zero. To estimate SNP-based heritability 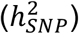, individual-level genotype data were used to derive empirical kinship matrices using KING (version 2.7). 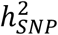 was then calculated using the QuantHer method in LDAK (version 4.9),^42^ which is suitable for the complex Amish pedigrees. The same covariates as above were controlled for in all models. We first estimated the heritability of biomarkers in the entire Amish cohort (All-Amish group). For sensitivity analysis, we repeated the calculation of 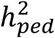 and 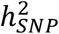 in two subgroups: individuals who were cognitively unimpaired (CU-only group) and those who did not carry any APOE ε4 alleles (non-*APOE* ε4 group).

## 3. Results

Table 1 provides a summary of the sample size, number of contributing sub-pedigrees, and key demographic characteristics within each biomarker-specific dataset, including age, sex, and the presence of *APOE* ε4 allele. The mean age of participants ranges from 80.6 to 82.1 years, reflecting a demographic representative of older populations typically studied in AD research. There’s a slightly higher percentage of females within each dataset, while the proportion of participants carrying the *APOE* ε4 allele varies between 22.2% and 26.2%.

**Table 1.**
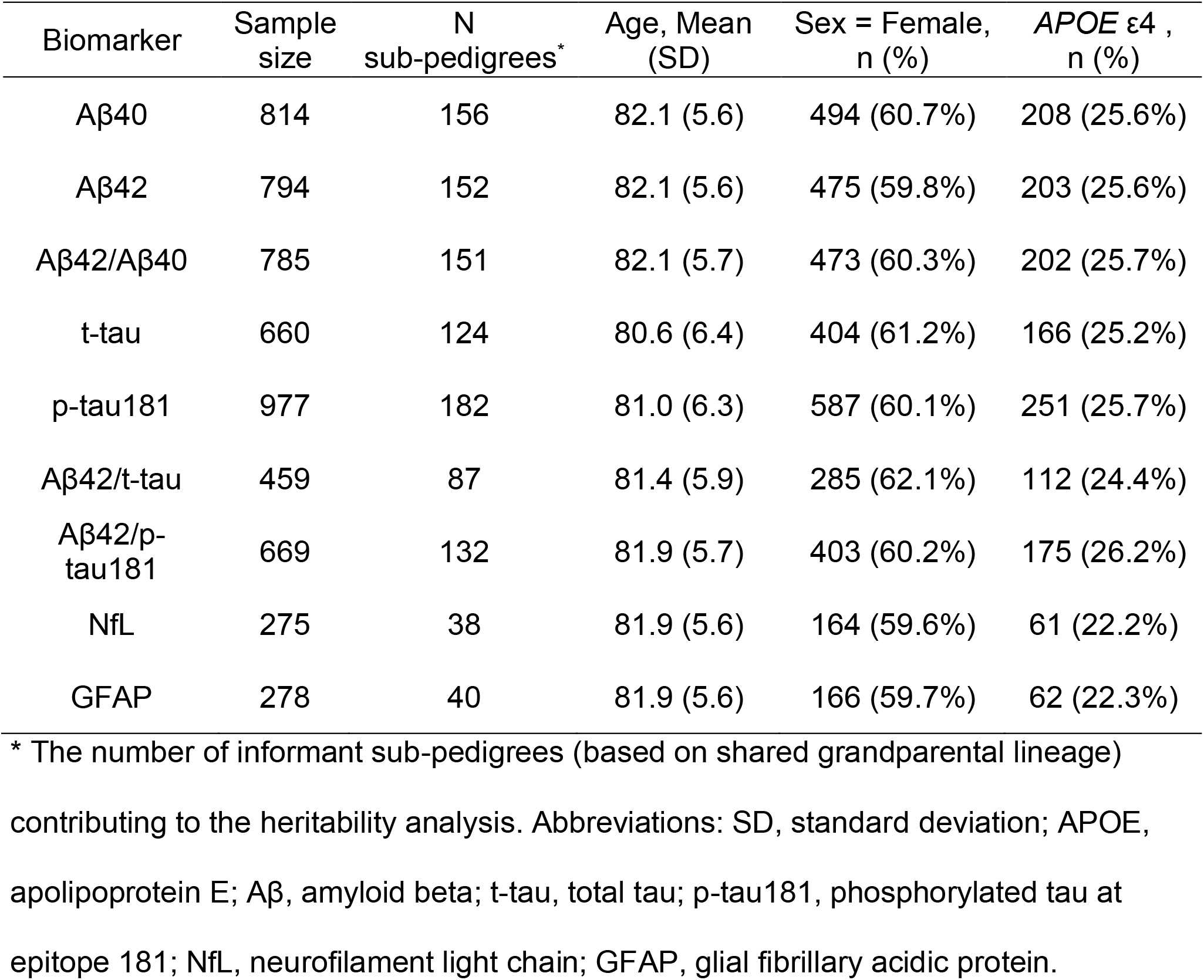
Profile of biomarker-specific datasets in the Amish.

| Biomarker | Sample size | N sub-pedigrees* | Age, Mean (SD) | Sex = Female, n (%) | APOE ε4 , n (%) |
| --- | --- | --- | --- | --- | --- |
| Aβ40 | 814 | 156 | 82.1 (5.6) | 494 (60.7%) | 208 (25.6%) |
| Aβ42 | 794 | 152 | 82.1 (5.6) | 475 (59.8%) | 203 (25.6%) |
| Aβ42/Aβ40 | 785 | 151 | 82.1 (5.7) | 473 (60.3%) | 202 (25.7%) |
| t-tau | 660 | 124 | 80.6 (6.4) | 404 (61.2%) | 166 (25.2%) |
| p-tau181 | 977 | 182 | 81.0 (6.3) | 587 (60.1%) | 251 (25.7%) |
| Aβ42/t-tau | 459 | 87 | 81.4 (5.9) | 285 (62.1%) | 112 (24.4%) |
| Aβ42/p-tau181 | 669 | 132 | 81.9 (5.7) | 403 (60.2%) | 175 (26.2%) |
| NfL | 275 | 38 | 81.9 (5.6) | 164 (59.6%) | 61 (22.2%) |
| GFAP | 278 | 40 | 81.9 (5.6) | 166 (59.7%) | 62 (22.3%) |

\* The number of informant sub-pedigrees (based on shared grandparental lineage) contributing to the heritability analysis. Abbreviations: SD, standard deviation; APOE, apolipoprotein E; Aβ, amyloid beta; t-tau, total tau; p-tau181, phosphorylated tau at epitope 181; NfL, neurofilament light chain; GFAP, glial fibrillary acidic protein.

As shown in Figure 1, no significant differences were found in plasma amyloid biomarkers, including Aβ40, Aβ42, and Aβ42/Aβ40 ratio, across diagnostic groups. Similarly, plasma t-tau did not significantly differ between groups. However, after adjusting for multiple comparisons, individuals in the AD group had significantly higher plasma p-tau181 levels than CU group (2.1 ± 0.9 pg/mL vs. 1.9 ± 0.8 pg/mL, *P*_*Bonferroni*_ = 0.04). For the plasma Aβ42/p-tau181 ratio, individuals with AD had significantly lower mean values (3.6 ± 2.6) compared to those with CINAD (5.5 ± 22.9, *P*_*Bonferroni*_ = 0.02) and CU individuals (4.8 ± 2.9, *P*_*Bonferroni*_ = 0.01). No significant differences were noted in the Aβ42/t-tau ratio between groups. Compared to the CU group, those in the AD group demonstrated significantly elevated levels of plasma Nfl (44.7 ± 15.3 pg/mL vs. 29.4 ± 15.1 pg/mL, *P*_*Bonferroni*_ = 0.01) and GFAP (171.1 ± 60.5 pg/mL vs. 130.8 ± 57.4 pg/mL, *P*_*Bonferroni*_ = 0.01).

### 3.1 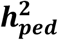 and 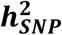 among all Amish individuals

Table 2 presents the heritability estimates for key AD-related plasma biomarkers in the All-Amish group. Overall, the heritability analysis revealed notable genetic influences across various measures, with pedigree-based heritability generally exceeding SNP-based estimates. Among all evaluated biomarkers, p-tau181 exhibited the highest heritability, with both pedigree-based (36.6%) and SNP-based (28.7%) estimates reaching statistical significance (*p* < 0.001). The heritability for other tau-related biomarkers (t-tau, Aβ42/t-tau, and Aβ42/p-tau181) was less pronounced. Amyloid β biomarkers (Aβ40, Aβ42, and Aβ42/Aβ40) showed moderate pedigree-based heritability ranging from 22.7% to 30.0% (*p* ≤ 0.007), with slightly lower but significant SNP-based heritability estimates (17.9% - 22.4%, *p* ≤ 0.034). In contrast, plasma NfL and GFAP presented lower, non-significant heritability estimates in both pedigree-based (11.1% - 21.1%, p ≥ 0.207) and SNP-based analyses (6.7% - 16.3%, *p* ≥ 0.337), suggesting weaker genetic influences on these markers for neuronal injury and neuroinflammation.

**Table 2.** Heritability of plasma biomarkers in the All-Amish group.

| | Sample<br>Size | $h^2_{ped}$ | | | $h^2_{SNP}$ | | |
| --- | --- | --- | --- | --- | --- | --- | --- |
|  |  | % | (SE) | *p* | % | (SE) | *p* |
| Aβ40 | 814 | 29.7% | (7.9%) | < 0.001 | 20.2% | (3.8%) | < 0.001 |
| Aβ42 | 794 | 22.7% | (8.9%) | 0.007 | 17.9% | (6.6%) | 0.034 |
| Aβ42/Aβ40 | 785 | 30.0% | (10.8%) | 0.002 | 22.4% | (6.7%) | 0.014 |
| t-tau | 660 | 31.0% | (12.8%) | 0.006 | 19.8% | (7.3%) | 0.045 |
| p-tau181 | 977 | 36.6% | (7.3%) | < 0.001 | 28.7% | (4.5%) | < 0.001 |
| Aβ42/t-tau | 459 | 30.2% | (18.9%) | 0.052 | 23.3% | (11.0%) | 0.140 |
| Aβ42/p-tau181 | 669 | 13.8% | (10.9%) | 0.093 | 12.0% | (7.6%) | 0.190 |
| NfL | 275 | 21.1% | (27.6%) | 0.207 | 16.3% | (9.3%) | 0.337 |
| GFAP | 278 | 11.1% | (19.1%) | 0.348 | 6.7% | (10.9%) | 0.565 |

Abbreviations: $h^2_{ped}$ , pedigree-based heritability; $h^2_{SNP}$ , SNP-based heritability; SE, standard error; Aβ, amyloid beta; t-tau, total tau; p-tau181, phosphorylated tau at epitope 181; NfL, neurofilament light chain; GFAP, glial fibrillary acidic protein.

For the covariates, age consistently showed a statistically significant impact (*p* < 0.05) on nearly all biomarkers, except for the Aβ42/t-tau and Aβ42/p-tau181 ratios. Sex significantly influenced Aβ40 (*p* = 0.02) and GFAP (*p* < 0.001). The presence of *APOE* ε4 alleles was strongly associated with Aβ42/Aβ40, p-tau181, and Aβ42/p-tau181 (*p* < 0.001). Additionally, variability introduced by the mixed use of biomarker kits (e.g., Simoa™ Neurology 3-Plex vs. 4-Plex assays) likely contributed to significant effects of biomarker batches on three amyloid β biomarkers, as well as p-tau181 and Aβ42/p-tau181 (*p* < 0.001).

### 3.1 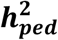 and 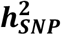 among CU individuals

To further investigate the heritability of plasma biomarkers in the elderly population with normal cognition, we calculated 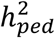 and 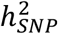 in the CU-only group (Table 3). Due to the limited sample size for NfL and GFAP (n < 170), we focused our analysis on amyloid β and tau-related biomarkers. In the CU-only group, the heritability of Aβ40 showed a modest increase compared to the estimates derived from the All-Amish group. Specifically, 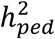 was 36.3% (*p* = 0.005), while 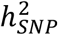 was 31.7% (*p* = 0.008), both remaining statistically significant. When comparing Aβ42 heritability in the All-Amish group, the CU-only group showed a slightly higher 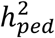 of 26.1% (*p* = 0.037) but a lower 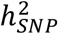 of 15.4% (*p* = 0.185). For Aβ42/Aβ40 ratio, however, both 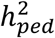 and 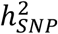 decreased and became statistically insignificant (*p* = 0.139 and 0.514, respectively). The heritability estimates for p-tau181 in the CU-only group were similar to those observed in the All-Amish group 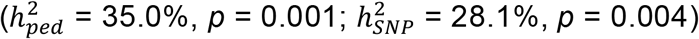. However, t-tau did not appear to be as heritable, with a 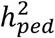 of 1.9% (*p* = 0.451) and a 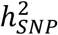 of 0.90% (*p* = 0.514). The heritability of Aβ42/t-tau and Aβ42/p-tau181 are not statistically significant in the CU-only group, and the estimates for both biomarkers were lower than those derived from the All-Amish group.

**Table 3.** Heritability of plasma biomarkers in the CU-only group.

| | Sample Size | $h^2_{ped}$ | | | $h^2_{SNP}$ | | |
| --- | --- | --- | --- | --- | --- | --- | --- |
|  |  | % | (SE) | *p* | % | (SE) | *p* |
| Aβ40 | 548 | 36.3% | (14.2%) | 0.005 | 31.7% | (7.9%) | 0.008 |
| Aβ42 | 533 | 26.1% | (14.8%) | 0.037 | 15.4% | (9.2%) | 0.185 |
| Aβ42/Aβ40 | 528 | 18.6% | (17.3%) | 0.139 | 9.1% | (11.5%) | 0.514 |
| t-tau | 445 | 1.9% | (15.1%) | 0.451 | 0.9% | (13.9%) | 0.950 |
| p-tau181 | 645 | 35.0% | (12.4%) | 0.001 | 28.1% | (6.6%) | 0.004 |
| Aβ42/t-tau | 325 | 28.1% | (22.8%) | 0.108 | 19.9% | (13.8%) | 0.307 |
| Aβ42/p-tau181 | 466 | 8.2% | (13.7%) | 0.301 | 7.4% | (9.7%) | 0.500 |

Abbreviations: CU, cognitively unimpaired; $h^2_{ped}$ , pedigree-based heritability; $h^2_{SNP}$ , SNP-based heritability; SE, standard error; Aβ, amyloid beta; t-tau, total tau; p-tau181, phosphorylated tau at epitope 181.

Age remained a statistically significant covariate in explaining the phenotypic variance of Aβ40, Aβ42, t-tau, and p-tau181 (*p* < 0.05). Sex demonstrated significant associations with Aβ40 (*p* = 0.02), t-tau (*p* = 0.03), and Aβ42/t-tau ratio (*p* = 0.01). The presence of *APOE* ε4 alleles continued to show a significant influence on Aβ42/Aβ40, p-tau181, and Aβ42/p-tau181 (*p* < 0.05). Similarly, the effects of study center and biomarker batches on amyloid β biomarkers, p-tau181, and Aβ42/p-tau181 remained statistically significant (*p* < 0.05).

### 3.3 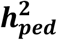 and 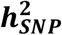 by *APOE* ε4 carrier status

Notably, only 22% - 26% of individuals across the biomarker-specific datasets carried an *APOE* ε4 allele. Due to the smaller sample sizes for NfL and GFAP, robust heritability estimates were feasible only among *APOE* ε4 non-carriers (Table 4). Heritability estimates were statistically significant for most amyloid β biomarkers, with the exception of 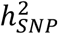 for Aβ42/Aβ40 ratio. In the non-*APOE* ε4 group, plasma Aβ42 had a 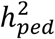 of 28.2% (*p* = 0.005) and a 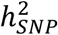 of 23.7% (*p* = 0.021), followed by slightly lower estimates for Aβ40 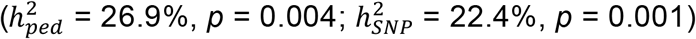 and Aβ42/Aβ40 ratio (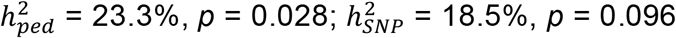). Compared to the heritability in the All-Amish group, the analysis of plasma t-tau yielded a comparable 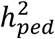 of 32.7% (*p* = 0.013) and a slightly higher 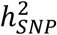 of 24.0% (*p* = 0.051). Plasma p-tau181 had the highest heritability among *APOE* ε4 non-carriers, with a 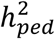 of 43.1% (*p* < 0.001) and a 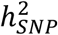 of 32.2% (*p* < 0.001). The 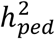 of Aβ42/t-tau and Aβ42/p-tau181 were significant, with values of 37.1% (*p* = 0.035) and 27.1% (*p* = 0.016), respectively. However, the 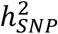 for these two ratios were no longer statistically significant, estimated at 29.6% (*p* = 0.11) for the Aβ42/t-tau ratio and 2.6% (*p* = 0.85) for the Aβ42/p-tau181 ratio.

**Table 4.** Heritability of plasma biomarkers in the non-*APOE* ε4 group.

| | Sample<br>Size | $h^2_{ped}$ | | | $h^2_{SNP}$ | | |
| --- | --- | --- | --- | --- | --- | --- | --- |
|  |  | % | (SE) | *p* | % | (SE) | *p* |
| Aβ40 | 606 | 26.9% | (5.9%) | 0.004 | 22.4% | (4.8%) | 0.001 |
| Aβ42 | 591 | 28.2% | (5.6%) | 0.005 | 23.7% | (7.5%) | 0.021 |
| Aβ42/Aβ40 | 583 | 23.3% | (9.9%) | 0.028 | 18.5% | (8.5%) | 0.096 |
| t-tau | 494 | 32.7% | (11.2%) | 0.013 | 24.0% | (8.7%) | 0.051 |
| p-tau181 | 726 | 43.1% | (10.1%) | < 0.001 | 32.2% | (5.3%) | < 0.001 |
| Aβ42/t-tau | 347 | 37.1% | (12.4%) | 0.035 | 29.6% | (12.2%) | 0.110 |
| Aβ42/p-tau181 | 494 | 27.1% | (14.1%) | 0.016 | 2.6% | (3.5%) | 0.850 |

Abbreviations: *APOE*, apolipoprotein E; $h^2_{ped}$ , pedigree-based heritability; $h^2_{SNP}$ , SNP- based heritability; SE, standard error; Aβ, amyloid beta; t-tau, total tau; p-tau181, phosphorylated tau at epitope 181.

Age was a statistically significant covariate in all heritability analyses (*p* < 0.05), except for the Aβ42/t-tau ratio. Sex showed a significant association only with Aβ40 (*p* = 0.03) and Aβ42 (*p* = 0.03). Consistent with previous findings, study center, and biomarker batch effects remained significant covariates in explaining the variances of amyloid β biomarkers, p-tau181, and the Aβ42/p-tau181 ratio (*p* < 0.001).

## 4. Discussion

We assessed the heritability of plasma Aβ40, Aβ42, Aβ42/Aβ40, t-tau, p-tau181, Aβ42/t-tau, Aβ42/p-tau181, NfL, and GFAP levels in the Midwestern Amish. For each biomarker, we calculated both the pedigree-based heritability and SNP-based heritability. In the All-Amish group, p-tau181 showed the highest heritability 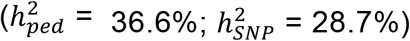, while GFAP exhibited the lowest 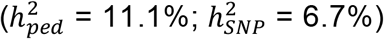. Overall, our findings are consistent with heritability estimates in previous studies,^25–27^ highlighting the substantial genetic contribution to the variation of AD-related plasma biomarkers. To our knowledge, this is the first study to comprehensively estimate both pedigree-based and SNP-based heritability of AD-related plasma biomarkers within a family-based design.

### 4.1 Pedigree-based vs. SNP-based heritability

We observed consistently higher 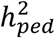 than 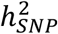, which likely reflects the contribution of more complex genetic models and/or rare variants not detected by SNP-based approaches.^43–46^ As a genetically isolated population, the Amish may have an enrichment of rare genetic variants due to the founder effect and limited gene flow.^37^ Rare variants can play a significant role in trait variability, but may not be captured by SNP arrays, even using imputation. In contrast, the pedigree-based approach considers the overall genetic relatedness and is more likely to capture the influence of rare variants as well as other inherited genomic features, such as structural variants and epistatic Interactions, leading to higher heritability estimates.^47^ Moreover, the Amish had relatively uniform shared environmental factors such as education, diets, and health behaviors.^48,49^ This homogeneity may reduce environmental variance, making variation explained by genetic components more apparent in pedigree-based analysis.

### 4.1 Heritability of plasma amyloid β biomarkers

The 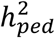 of amyloid β biomarkers (Aβ40, Aβ42, and Aβ42/Aβ40) ranged from 22.7% to 30.0%, and the 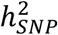 estimates were between 17.9% and 22.4%. When the analyses were restricted to the CU-only and non-*APOE* ε4 groups, both 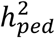 and 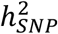 for amyloid β biomarkers fluctuated due to the changes in sample sizes and pedigree structures, but overall, our heritability estimates of individual Aβ40 and Aβ42 levels were lower than previously reported values.^25,27^ However, the plasma Aβ42/Aβ40 ratio was significantly heritable, in contrast to recent studies that found much lower heritability for this ratio.^26,27^ The Aβ42/Aβ40 ratio, adjusting for inter-individual differences in total Aβ production, often shows a higher concordance with brain amyloidosis.^50^ The ratio shares many of the same loci that affect individual Aβ40 or Aβ42 levels.^51,52^ However, some variants may preferentially alter the relative production or clearance of Aβ40 vs. Aβ42,^51^ causing this ratio to have partially distinct genetic architecture compared to either peptide alone. Such complexity underscores the need for further investigation into the genetic pathways underlying amyloid β biomarkers.

### 4.2 Heritability of plasma tau biomarkers

Plasma p-tau181 levels were significantly higher in the AD group compared to the CU group (*p* = 0.04), and p-tau181 demonstrated the strongest heritability with a 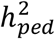 of 36.6% and 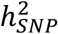 of 28.7% (*p* < 0.001). The estimates remained significant in the CU-only and non-*APOE* ε4 groups. Elevated p-tau181 levels are strongly associated with the disease onset, diagnosis, and progression,^53^ and the substantial heritable component of p-tau181 makes this marker a key candidate for identifying genetic loci linked to AD risk. A recent GWAS has shown that AD risk loci outside of *APOE* are primarily affecting plasma p-tau181.^52^ The 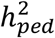 and 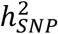 for plasma t-tau levels were comparable between the All-Amish and non-*APOE* ε4 groups. However, plasma t-tau did not appear to be heritable in the CU-only group, with a 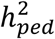 of 1.9% (*p* = 0.45) and a 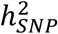 of 0.9% (*p* = 0.95). This could be due to the constrained phenotypic variance in this subgroup. Moreover, pathological changes in t-tau are not specific to AD, and previous studies have demonstrated plasma t-tau levels remain relatively low in cognitively normal individuals, and tau elevation is more pronounced in later stages of AD.^54^

### 4.4 Heritability of composite biomarkers

We evaluated the heritability of composite biomarkers, Aβ42/t-tau and Aβ42/p-tau181 ratios, to integrate implications from both pathologies. Although the Aβ42/t-tau ratio did not differ significantly between diagnostic groups, plasma Aβ42/t-tau appears to be a heritable trait with a 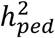 of 30.2% (*p* = 0.05) and a 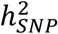 of 23.3% (*p* = 0.14) in the All-Amish group. Together with the results in the CU-only and non-*APOE* ε4 groups, the Aβ42/t-tau ratio demonstrated heritability estimates comparable to or exceeding those of individual Aβ42 and t-tau levels, suggesting that composite biomarkers may uncover additional genetic effects not apparent in single-marker analyses.

It’s worth noting that the Aβ42/p-tau181 ratio may be a superior measure in classifying AD in the Amish. The AD group had significantly lower values of Aβ42/p-tau181 ratio compared to individuals with CINAD (*p* = 0.02) and CU individuals (*p* = 0.01), in line with a previous study showing higher power of Aβ42/p-tau181 ratio differentiating AD over other neurodegenerative conditions.^55^ However, the heritability of the Aβ42/p-tau181 ratio was relatively low and not statistically significant. The decline in the heritability estimates of plasma Aβ42/p-tau181 ratio could stem from complex genetic and environmental factors. For example, there are distinct genetic pathways that influence Aβ42 and p-tau181^52,56^ and may not synergize, leading to attenuated heritability for the composite measure. From a statistics perspective, ratios alone do not account for variation in either measure, and calculating ratios may amplify measurement errors inherent in each biomarker assay, leading to inaccurate estimation heritability.^59^

### 4.5 Heritability of plasma NfL and GFAP

This study is among the first to estimate the heritability of plasma NfL and GFAP using a family-based design. NfL and GFAP are key indicators of neuronal damage and neuroinflammation in various neurodegenerative disorders.^81^ Individuals with AD have elevated concentrations of NfL long before the clinical onset of symptoms; furthermore, the greater the increase in NfL, the more rapidly the clinical illness progresses.^60,61^ Recent data also suggests increased concentrations of GFAP in individuals with AD.^22,62^ We found significantly elevated levels of plasma Nfl and GFAP in Amish individuals with AD compared to CU individuals (*p* = 0.01). Recent twin studies reported the heritability of plasma NfL to be 42% - 43% and 60% for plasma GFAP.^26,27^ However, our results suggest a lower heritability, maximally 21.1% and 11.1% for NfL and GFAP, respectively. Several factors likely contribute to these discrepancies. First, the small sample sizes of NfL and GFAP in the Amish reduced the precision and power to detect genetic effects. Second, while twin studies can account for both additive and non-additive genetic effects in estimating heritability, they often do not consider the impact of shared environmental factors, which may lead to an overestimation of heritability.^63,64^ In contrast, pedigree-based and SNP-based methods focus on additive effects, resulting in lower estimates if non-additive effects contribute significantly. Finally, numerous studies have found elevated serum NfL and/or GFAP levels associated with environmental risk factors, including smoking, alcohol consumption, and air pollution.^65–69^ The Amish experience reduced exposure to these environmental risks, which may lower the overall variability of plasma NfL and GFAP and make it harder to identify genetic contributions.

### 4.6 Role of age in heritability analyses

Age consistently emerged as a significant covariate in all heritability models, highlighting its critical role in biomarker variability. Aging influences multiple biological systems, including the blood-brain barrier and neuroinflammatory pathways, affecting the production, clearance, and stability of fluid biomarkers.^70^ Additionally, genetic influences on biomarker levels may vary across the lifespan, with some effects becoming more pronounced or diminishing over time.^71^ It is essential for future studies to adjust for age in the genetic analysis to disentangle genetic contributions from age-related changes and ensure more accurate heritability estimates.

### 4.7 Study limitations

This study should be interpreted in the context of several limitations. First, the Amish are founder populations with reduced genetic diversity and relatively uniform environmental exposures. This may limit the ability to detect environmental influences on biomarker levels and thus generalize findings to broader populations. Second, SNP-based heritability relies on common variants and the underlying reference population used for imputation. In this study, we incorporate 6,000 more variants for genotyping, including over 1,100 Amish-specific novel variants, to optimize the imputed genotype data used for analysis.^31,32,36,37^ However, rare variants that are more enriched in the Amish and contribute to biomarker variability may remain undetected. Third, some of the biomarker data in this study were obtained from historically collected plasma samples and measured using a mix of Simoa™ assays. Composite biomarkers such as ratios may amplify any technical differences. Fourth, the AD diagnosis relies mostly on medical history and detailed cognitive measurements, as the Amish do not have imaging or CSF data available. We adjusted for non-genetic factors, such as age and sex, but other factors (e.g., comorbidities) that may confound heritability analyses were unavailable in this study.

## 5. Conclusion

Plasma biomarkers, as quantitative endophenotypes of AD, offer a window into disease liability. Understanding the genetic determinants of these biomarkers is critical for elucidating biological pathways involved in AD pathogenesis, improving risk stratification, and enabling earlier, potentially more effective interventions. To our knowledge, this is the first study leveraging both pedigree and genotype data in a family-based design to estimate the heritability of core AD-related biomarkers. The pedigree-based heritability for our suite of biomarkers ranged from 11.1% to 36.6%, and SNP-based heritability ranged from 6.7% to 28.7% among all Amish individuals. These findings confirmed that plasma amyloid β and tau biomarkers have substantial heritable components, and this study emphasizes the importance of subgroup analyses in heritability estimation, as genetic influences on biomarkers may vary significantly across different study groups and disease stages. Future studies are needed to incorporate additional genetic (e.g., rare variants, epigenetics) and environmental factors to provide deeper insights into the determinants of plasma biomarkers associated with AD.

## Data Availability

All data produced in the present study are available upon reasonable request to the authors.

## Acknowledgements

We thank the Amish families for their willingness to participate in our study. This research was supported by the National Institutes of Health and National Institute on Aging (grants AG019085, AG019726, and AG058066).

## Conflicts

The authors declare no conflicts of interest.

## Funding Sources

National institutes of Health and National Institute on Aging (grants AG019085, AG019726, and AG058066).

## Consent Statement

All study procedures were approved by the Institutional Review Board at the Case Western Reserve University and University of Miami. Written informed consent was obtained from the participant or legal guardian. For the sake of confidentiality, clinical and genetic data were de-identified according to the study protocol.

**Supplementary Figure 1.**
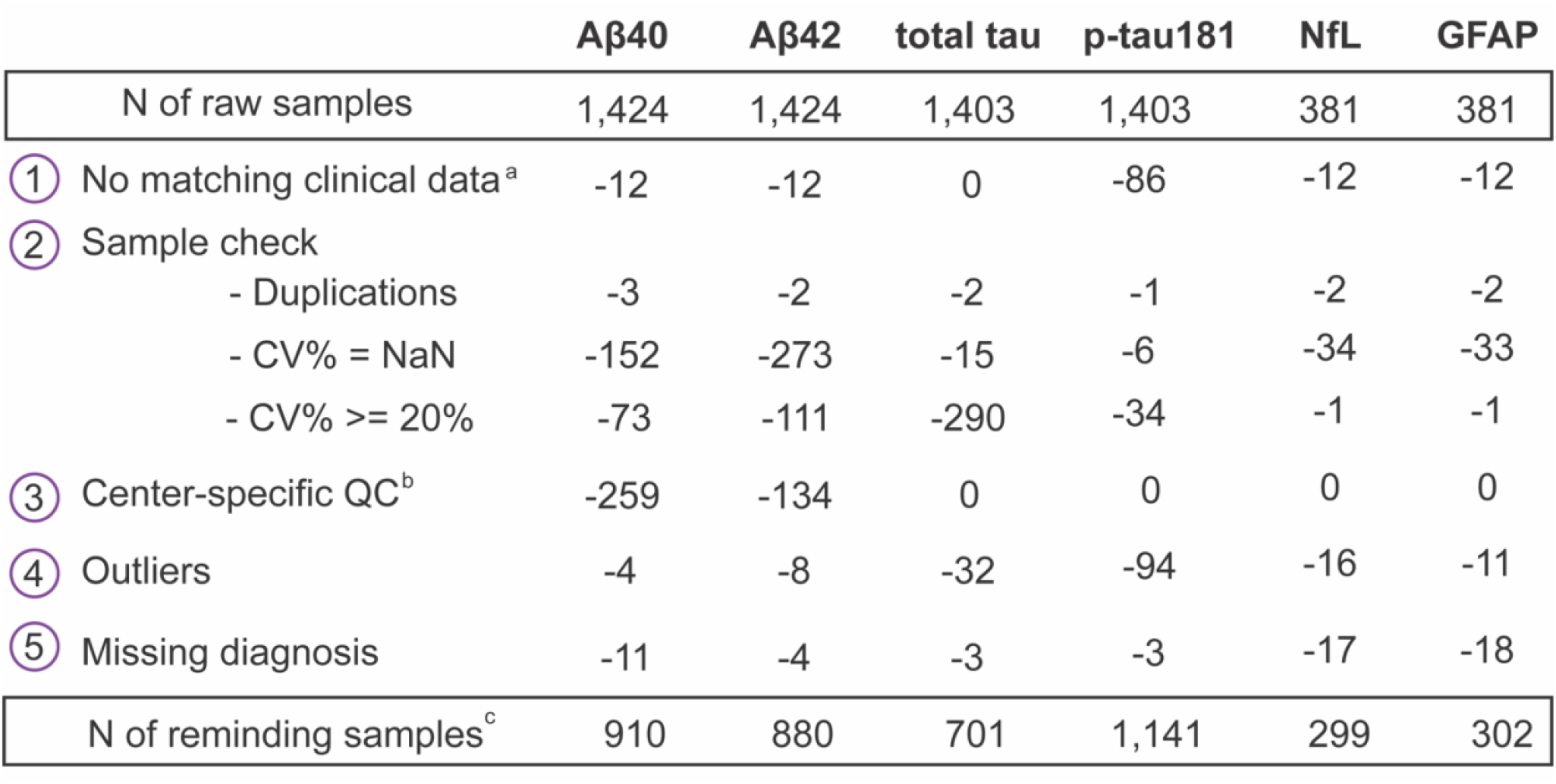
Biomarker-level quality control a. Samples had no matching clinical data (i.e., no consensus diagnosis or not Amish). b. Aβ40 and Aβ42 samples before 2014 from Ohio were excluded. c. Samples passed QC include one-time and repeated measurements on the same individual. Abbreviations: QC, quality control; CV, Coefficient of variation percentages; Aβ, amyloid beta; p-tau181, phosphorylated tau at epitope 181; NfL, neurofilament light chain; GFAP, glial fibrillary acidic protein.

## Notes

### Competing Interest Statement

The authors have declared no competing interest.

### Funding Statement

This research was funded by the National Institutes of Health and National Institute on Aging (grants AG019085, AG019726, and AG058066).

### Author Declarations

The Institutional Review Board of Case Western Reserve University and the University of Miami gave ethical approval for this work.

### Summary of Updates

I identified a typo in the manuscript after submission, this revision is to correct it.

## Reference

1. Jansen IE, Savage JE, Watanabe K, et al. Genome-wide meta-analysis identifies new loci and functional pathways influencing Alzheimer’s disease risk. Nat Genet. 2019/03/01 2019;51(3):404–413. doi:10.1038/s41588-018-0311-9

2. Kunkle BW, Grenier-Boley B, Sims R, et al. Genetic meta-analysis of diagnosed Alzheimer’s disease identifies new risk loci and implicates Aβ, tau, immunity and lipid processing. Nat Genet. Mar 2019;51(3):414–430. doi:10.1038/s41588-019-0358-2

3. Naj AC, Leonenko G, Jian X, et al. Genome-Wide Meta-Analysis of Late-Onset Alzheimer’s Disease Using Rare Variant Imputation in 65,602 Subjects Identifies Novel Rare Variant Locus <em>NCK2</em>: The International Genomics of Alzheimer’s Project (IGAP). medRxiv. 2021:2021.03.14.21253553. doi:10.1101/2021.03.14.21253553

4. Schwartzentruber J, Cooper S, Liu JZ, et al. Genome-wide meta-analysis, fine-mapping and integrative prioritization implicate new Alzheimer’s disease risk genes. Nat Genet. 2021/03/01 2021;53(3):392–402. doi:10.1038/s41588-020-00776-w

5. Bellenguez C, Küçükali F, Jansen IE, et al. New insights into the genetic etiology of Alzheimer’s disease and related dementias. Nat Genet. 2022/04/01 2022;54(4):412–436. doi:10.1038/s41588-022-01024-z

6. Pedersen NL, Gatz M, Berg S, Johansson B. How heritable is Alzheimer’s disease late in life? Findings from Swedish twins. Annals of Neurology. 2004;55(2):180–185. doi:10.1002/ana.10999

7. Gatz M, Pedersen NL, Berg S, et al. Heritability for Alzheimer’s disease: the study of dementia in Swedish twins. J Gerontol A Biol Sci Med Sci. Mar 1997;52(2):M117–25. doi:10.1093/gerona/52a.2.m117

8. Bergem AL, Engedal K, Kringlen E. The role of heredity in late-onset Alzheimer disease and vascular dementia. A twin study. Arch Gen Psychiatry. Mar 1997;54(3):264–70. doi:10.1001/archpsyc.1997.01830150090013

9. Ridge PG, Mukherjee S, Crane PK, Kauwe JSK, Alzheimer’s Disease Genetics C. Alzheimer’s Disease: Analyzing the Missing Heritability. PLOS ONE. 2013;8(11):e79771. doi:10.1371/journal.pone.0079771

10. Lee SH, Harold D, Nyholt DR, et al. Estimation and partitioning of polygenic variation captured by common SNPs for Alzheimer’s disease, multiple sclerosis and endometriosis. Human Molecular Genetics. 2012;22(4):832–841. doi:10.1093/hmg/dds491

11. Génin E. Missing heritability of complex diseases: case solved? Human Genetics. 2020/01/01 2020;139(1):103–113. doi:10.1007/s00439-019-02034-4

12. Migliore L, Coppedè F. Gene–environment interactions in Alzheimer disease: the emerging role of epigenetics. Nature Reviews Neurology. 2022/11/01 2022;18(11):643–660. doi:10.1038/s41582-022-00714-w

13. Dunn AR, O’Connell KMS, Kaczorowski CC. Gene-by-environment interactions in Alzheimer’s disease and Parkinson’s disease. Neuroscience & Biobehavioral Reviews. 2019/08/01/ 2019;103:73–80. doi:10.1016/j.neubiorev.2019.06.018

14. Zhang Q, Liu J, Liu H, Ao L, Xi Y, Chen D. Genome-wide epistasis analysis reveals gene–gene interaction network on an intermediate endophenotype P-tau/Aβ42 ratio in ADNI cohort. Scientific Reports. 2024/02/17 2024;14(1):3984. doi:10.1038/s41598-024-54541-8

15. Hohman TJ, Bush WS, Jiang L, et al. Discovery of gene-gene interactions across multiple independent data sets of late onset Alzheimer disease from the Alzheimer Disease Genetics Consortium. Neurobiology of Aging. 2016/02/01/ 2016;38:141–150. doi:10.1016/j.neurobiolaging.2015.10.031

16. Kunkle BW, Schmidt M, Klein H-U, et al. Novel Alzheimer Disease Risk Loci and Pathways in African American Individuals Using the African Genome Resources Panel: A Meta-analysis. JAMA Neurology. 2021;78(1):102–113. doi:10.1001/jamaneurol.2020.3536

17. Beach TG, Monsell SE, Phillips LE, Kukull W. Accuracy of the clinical diagnosis of Alzheimer disease at National Institute on Aging Alzheimer Disease Centers, 2005-2010. J Neuropathol Exp Neurol. Apr 2012;71(4):266–73. doi:10.1097/NEN.0b013e31824b211b

18. Gao S, Wang T, Han Z, et al. Interpretation of 10 years of Alzheimer’s disease genetic findings in the perspective of statistical heterogeneity. Briefings in Bioinformatics. 2024;25(3) doi:10.1093/bib/bbae140

19. Hardy J, Allsop D. Amyloid deposition as the central event in the aetiology of Alzheimer’s disease. Trends in pharmacological sciences. 1991;12:383–388.

20. Braak H, Braak E. Neuropathological stageing of Alzheimer-related changes. Acta Neuropathol. 1991;82(4):239–59. doi:10.1007/bf00308809

21. Khalil M, Teunissen CE, Otto M, et al. Neurofilaments as biomarkers in neurological disorders. Nature Reviews Neurology. 2018;14(10):577–589.

22. Pereira JB, Janelidze S, Smith R, et al. Plasma GFAP is an early marker of amyloid-β but not tau pathology in Alzheimer’s disease. Brain. Dec 16 2021;144(11):3505–3516. doi:10.1093/brain/awab223

23. Blennow K. Phenotyping Alzheimer’s disease with blood tests. Science. 2021;373(6555):626–628. doi:doi:10.1126/science.abi5208

24. Polacchini A, Metelli G, Francavilla R, et al. A method for reproducible measurements of serum BDNF: comparison of the performance of six commercial assays. Scientific Reports. 2015/12/10 2015;5(1):17989. doi:10.1038/srep17989

25. Ertekin-Taner N, Graff-Radford N, Younkin LH, et al. Heritability of plasma amyloid β in typical late-onset Alzheimer’s disease pedigrees. Genetic Epidemiology. 2001;21(1):19–30. doi:10.1002/gepi.1015

26. Rousset RZ, den Braber A, Verberk IMW, et al. Heritability of Alzheimer’s disease plasma biomarkers: A nuclear twin family design. Alzheimer’s & Dementia. 2024;n/a(n/a)doi:10.1002/alz.14269

27. Gillespie NA, Elman JA, McKenzie RE, et al. The heritability of blood-based biomarkers related to risk of Alzheimer’s disease in a population-based sample of early old-age men. Alzheimers Dement. Jan 2024;20(1):356–365. doi:10.1002/alz.13407

28. van der Walt JM, Scott WK, Slifer S, et al. Maternal lineages and Alzheimer disease risk in the Old Order Amish. Human genetics. 2005/10// 2005;118(1):115–122. doi:10.1007/s00439-005-0032-x

29. Hostetler JA. Amish society. JHU Press; 1993.

30. Agarwala R, Biesecker LG, Tomlin JF, Schäffer AA. Towards a complete North American Anabaptist genealogy: A systematic approach to merging partially overlapping genealogy resources. Am J Med Genet. Sep 10 1999;86(2):156–61. doi:10.1002/(sici)1096-8628(19990910)86:2<156::aid-ajmg13>3.0.co;2-5

31. Edwards DRV, Gilbert JR, Jiang L, et al. Successful aging shows linkage to chromosomes 6, 7, and 14 in the Amish. Annals of human genetics. 2011;75(4):516–528.

32. Edwards DR, Gilbert JR, Hicks JE, et al. Linkage and association of successful aging to the 6q25 region in large Amish kindreds. Age (Dordr). Aug 2013;35(4):1467–77. doi:10.1007/s11357-012-9447-1

33. Analyzer H-XAI. https://www.quanterix.com/instruments/simoa-hd-x-analyzer/

34. Consortium M-EGA. https://www.illumina.com/science/consortia/human-consortia/multi-ethnic-genotyping-consortium.html

35. Consortium GSA. https://www.illumina.com/science/consortia/human-consortia/global-screening-consortium.html

36. D’Aoust LN, Cummings AC, Laux R, et al. Examination of Candidate Exonic Variants for Association to Alzheimer Disease in the Amish. PLOS ONE. 2015;10(2):e0118043. doi:10.1371/journal.pone.0118043

37. Osterman MD, Song YE, Adams LD, et al. The genetic architecture of Alzheimer disease risk in the Ohio and Indiana Amish. Human Genetics and Genomics Advances. 2022/07/14/ 2022;3(3):100114. doi:10.1016/j.xhgg.2022.100114

38. Beecham GW, Bis J, Martin E, et al. The Alzheimer’s Disease Sequencing Project: study design and sample selection. Neurology Genetics. 2017;3(5)

39. Taliun D, Harris DN, Kessler MD, et al. Sequencing of 53,831 diverse genomes from the NHLBI TOPMed Program. Nature. 2021/02/01 2021;590(7845):290–299. doi:10.1038/s41586-021-03205-y

40. Main LR, Song YE, Lynn A, et al. Genetic analysis of cognitive preservation in the midwestern Amish reveals a novel locus on chromosome 2. Alzheimer’s & Dementia. 2024;20(11):7453–7464. doi:10.1002/alz.14045

41. Almasy L, Blangero J. Multipoint quantitative-trait linkage analysis in general pedigrees. The American Journal of Human Genetics. 1998;62(5):1198–1211.

42. Speed D, Evans DM. Estimating disease heritability from complex pedigrees allowing for ascertainment and covariates. The American Journal of Human Genetics. 2024;111(4):680–690. doi:10.1016/j.ajhg.2024.02.010

43. Manolio TA, Collins FS, Cox NJ, et al. Finding the missing heritability of complex diseases. Nature. Oct 8 2009;461(7265):747–53. doi:10.1038/nature08494

44. Yang J, Zeng J, Goddard ME, Wray NR, Visscher PM. Concepts, estimation and interpretation of SNP-based heritability. Nat Genet. 2017/09/01 2017;49(9):1304–1310. doi:10.1038/ng.3941

45. Visscher PM, Hill WG, Wray NR. Heritability in the genomics era — concepts and misconceptions. Nature Reviews Genetics. 2008/04/01 2008;9(4):255–266. doi:10.1038/nrg2322

46. Bochud M. Estimating Heritability from Nuclear Family and Pedigree Data. In: Elston RC, Satagopan JM, Sun S, eds. Statistical Human Genetics: Methods and Protocols. Humana Press; 2012:171–186.

47. Xia C, Amador C, Huffman J, et al. Pedigree- and SNP-Associated Genetics and Recent Environment are the Major Contributors to Anthropometric and Cardiometabolic Trait Variation. PLOS Genetics. 2016;12(2):e1005804. doi:10.1371/journal.pgen.1005804

48. Ramos J, Chowdhury AR, Caywood LJ, et al. Lower Levels of Education Are Associated with Cognitive Impairment in the Old Order Amish. J Alzheimers Dis. 2021;79(1):451–458. doi:10.3233/jad-200909

49. Mitchell BD, Schäffer AA, Pollin TI, et al. Mapping Genes in Isolated Populations: Lessons from the Old Order Amish. In: Duggirala R, Almasy L, Williams-Blangero S, Paul SFD, Kole C, eds. Genome Mapping and Genomics in Human and Non-Human Primates. Springer Berlin Heidelberg; 2015:141–153.

50. Schindler SE, Bollinger JG, Ovod V, et al. High-precision plasma β-amyloid 42/40 predicts current and future brain amyloidosis. Neurology. 2019;93(17):e1647–e1659.

51. Damotte V, van der Lee SJ, Chouraki V, et al. Plasma amyloid β levels are driven by genetic variants near APOE, BACE1, APP, PSEN2: A genome-wide association study in over 12,000 non-demented participants. Alzheimers Dement. Oct 2021;17(10):1663–1674. doi:10.1002/alz.12333

52. Bradley J, Gorijala P, Schindler SE, et al. Genetic architecture of plasma Alzheimer disease biomarkers. Hum Mol Genet. Jul 20 2023;32(15):2532–2543. doi:10.1093/hmg/ddad087

53. Janelidze S, Mattsson N, Palmqvist S, et al. Plasma P-tau181 in Alzheimer’s disease: relationship to other biomarkers, differential diagnosis, neuropathology and longitudinal progression to Alzheimer’s dementia. Nat Med. 2020/03/01 2020;26(3):379–386. doi:10.1038/s41591-020-0755-1

54. Mielke MM, Hagen CE, Wennberg AMV, et al. Association of Plasma Total Tau Level With Cognitive Decline and Risk of Mild Cognitive Impairment or Dementia in the Mayo Clinic Study on Aging. JAMA Neurology. 2017;74(9):1073–1080. doi:10.1001/jamaneurol.2017.1359

55. Vergallo A, Carlesi C, Pagni C, et al. A single center study: Aβ42/p-Tau181 CSF ratio to discriminate AD from FTD in clinical setting. Neurological Sciences. 2017/10/01 2017;38(10):1791–1797. doi:10.1007/s10072-017-3053-z

56. Stevenson-Hoare J, Heslegrave A, Leonenko G, et al. Plasma biomarkers and genetics in the diagnosis and prediction of Alzheimer’s disease. Brain. Feb 13 2023;146(2):690–699. doi:10.1093/brain/awac128

57. Martínez-Dubarbie F, Guerra-Ruiz A, López-García S, et al. Influence of Physiological Variables and Comorbidities on Plasma Aβ40, Aβ42, and p-tau181 Levels in Cognitively Unimpaired Individuals. International Journal of Molecular Sciences. 2024;25(3):1481.

58. Pan F, Lu Y, Huang Q, Xie F, Yang J, Guo Q. The potential impact of clinical factors on blood-based biomarkers for Alzheimer’s disease. Translational Neurodegeneration. 2023/08/18 2023;12(1):39. doi:10.1186/s40035-023-00371-z

59. White E. Measurement error in biomarkers: sources, assessment, and impact on studies. IARC Sci Publ. 2011;(163):143–61.

60. Clark C, Lewczuk P, Kornhuber J, et al. Plasma neurofilament light and phosphorylated tau 181 as biomarkers of Alzheimer’s disease pathology and clinical disease progression. Alzheimers Res Ther. Mar 25 2021;13(1):65. doi:10.1186/s13195-021-00805-8

61. Mattsson N, Cullen NC, Andreasson U, Zetterberg H, Blennow K. Association between longitudinal plasma neurofilament light and neurodegeneration in patients with Alzheimer disease. JAMA neurology. 2019;76(7):791–799.

62. Elahi FM, Casaletto KB, La Joie R, et al. Plasma biomarkers of astrocytic and neuronal dysfunction in early- and late-onset Alzheimer’s disease. Alzheimer’s & Dementia. 2020;16(4):681–695. doi:10.1016/j.jalz.2019.09.004

63. Rettew DC, Rebollo-Mesa I, Hudziak JJ, Willemsen G, Boomsma DI. Non-additive and additive genetic effects on extraversion in 3314 Dutch adolescent twins and their parents. Behav Genet. May 2008;38(3):223–33. doi:10.1007/s10519-008-9192-5

64. Dalmaijer ES. Twin studies with unmet assumptions are biased towards genetic heritability. bioRxiv. 2020:2020.08.27.270801. doi:10.1101/2020.08.27.270801

65. Zhu N, Zhu J, Lin S, Yu H, Cao C. Correlation analysis between smoke exposure and serum neurofilament light chain in adults: a cross-sectional study. BMC Public Health. Feb 2 2024;24(1):353. doi:10.1186/s12889-024-17811-8

66. Xue Y, Tran M, Diep YN, et al. Environmental aluminum oxide inducing neurodegeneration in human neurovascular unit with immunity. Scientific Reports. 2024/01/07 2024;14(1):744. doi:10.1038/s41598-024-51206-4

67. Li Y, Duan R, Gong Z, et al. Neurofilament Light Chain Is a Promising Biomarker in Alcohol Dependence. Original Research. Frontiers in Psychiatry. 2021-November-18 2021;12 doi:10.3389/fpsyt.2021.754969

68. Kang YJ, Tan HY, Lee CY, Cho H. An Air Particulate Pollutant Induces Neuroinflammation and Neurodegeneration in Human Brain Models. Adv Sci (Weinh). Nov 2021;8(21):e2101251. doi:10.1002/advs.202101251

69. Huf F, Bandiera S, Müller CB, et al. Comparative study on the effects of cigarette smoke exposure, ethanol consumption and association: Behavioral parameters, apoptosis, glial fibrillary acid protein and S100β immunoreactivity in different regions of the rat hippocampus. Alcohol. 2019/06/01/ 2019;77:101–112. doi:10.1016/j.alcohol.2018.08.009

70. Crimmins E, Vasunilashorn S, Kim JK, Alley D. Biomarkers related to aging in human populations. Adv Clin Chem. 2008;46:161–216. doi:10.1016/s0065-2423(08)00405-8

71. Winkler TW, Wiegrebe S, Herold JM, Stark KJ, Küchenhoff H, Heid IM. Genetic-by-age interaction analyses on complex traits in UK Biobank and their potential to identify effects on longitudinal trait change. Genome Biology. 2024/11/28 2024;25(1):300. doi:10.1186/s13059-024-03439-9

